# Decoding the role of transcriptomic clocks in the human prefrontal cortex

**DOI:** 10.1101/2023.04.19.23288765

**Authors:** José J. Martínez-Magaña, John H. Krystal, Matthew J. Girgenti, Diana L. Núnez-Ríos, Sheila T. Nagamatsu, Diego E. Andrade-Brito, Traumatic Stress Brain Research Group, Janitza L. Montalvo-Ortiz

**Affiliations:** Division of Human Genetics, Department of Psychiatry, Yale University School of Medicine, New Haven; National Center for PTSD, US Department of Veterans Affairs, West Haven, CT, USA; Psychiatry Service, VA Connecticut Health Care System, West Haven, CT, USA

**Keywords:** Biological clocks, aging, prefrontal cortex, transcriptomic clocks, deep learning

## Abstract

Aging is a complex process with interindividual variability, which can be measured by aging biological clocks. Aging clocks are machine-learning algorithms guided by biological information and associated with mortality risk and a wide range of health outcomes. One of these aging clocks are transcriptomic clocks, which uses gene expression data to predict biological age; however, their functional role is unknown. Here, we profiled two transcriptomic clocks (RNAAgeCalc and knowledge-based deep neural network clock) in a large dataset of human postmortem prefrontal cortex (PFC) samples. We identified that deep-learning transcriptomic clock outperforms RNAAgeCalc to predict transcriptomic age in the human PFC. We identified associations of transcriptomic clocks with psychiatric-related traits. Further, we applied system biology algorithms to identify common gene networks among both clocks and performed pathways enrichment analyses to assess its functionality and prioritize genes involved in the aging processes. Identified gene networks showed enrichment for diseases of signal transduction by growth factor receptors and second messenger pathways. We also observed enrichment of genome-wide signals of mental and physical health outcomes and identified genes previously associated with human brain aging. Our findings suggest a link between transcriptomic aging and health disorders, including psychiatric traits. Further, it reveals functional genes within the human PFC that may play an important role in aging and health risk.

## Main

Aging is a complex biological process that can manifest differently and at a different rate in each individual ^1–3^. The individual variability in biological aging can be measured, resulting in the development of aging clocks ^4–6^. Aging clocks are an excellent predictor of mortality and lifespan ^7–10^, as well as disease risk ^11,12^. One of the best-known aging clocks are epigenetic clocks ^13,14^, which are based on age-related alterations in DNA methylation at CpG sites. We and others have found a link between epigenetic clocks and psychiatric disorders ^15–18^. Age- associated biological changes can also occur at the RNA level ^19,20^. Recent studies of transcriptomic clocks ^21,22^ have shown associations with age-related disorders ^20^. These clocks have been generated using several mathematical models, including the elastic net model. However, such method is limited to a fixed set of genes for predicting age. Recently developed deep learning methods can overcome this limitation ^22^ by using the whole transcriptome to train a neural network to select the most precisely defined and informative set of genes.

The human prefrontal cortex (PFC) and other brain regions undergo molecular, structural, and functional changes during aging ^23–25^. Structural age-related alterations in the PFC in older individuals are well-established by neuroimaging studies ^26–32^. A meta-analysis of 3,880 individuals showed that older people have a reduction in the activation of subcortical and cortical brain regions compared with young adults measured by functional magnetic resonance during a cognitive task ^33^. Molecular studies evaluating brain aging have found that the PFC could age faster than other brain regions. For example, increased epigenetic age in the PFC is observed compared to the cerebellum ^34,35^. Further, accelerated epigenetic age in peripheral tissue has been associated with reduced cortical thickness ^36,37^. Recent work in aging clocks have shown that those trained in peripheral tissues do not have the same accuracy as those from the brain ^38^, with brain-trained clocks having a better chronological age prediction accuracy ^39^. While some work has evaluated epigenetic clocks in the brain, very few studies have examined transcriptomic clocks. Evaluating transcriptomic clocks is important given recent evidence showing age-associated transcriptomic changes in the PFC involving, for example, gene dysregulation of synaptic genes ^40–44^.

The present study aimed to profile a transcriptomic clock and to characterize its functional impact in four human PFC regions (**Fig. 1**) from a large cohort of 551 human postmortem brain samples. We highlight the advantage of deep learning methods to improve accuracy in predicting brain transcriptomic age and identify that both clocks have concordance at advanced ages. We replicated an association of the RNAAgeCalc with PTSD. Further, we comprehensively characterized co-expressed genes shared by both clocks and prioritized genes that could have an important role in aging within the human prefrontal cortex.

**Fig. 1.**
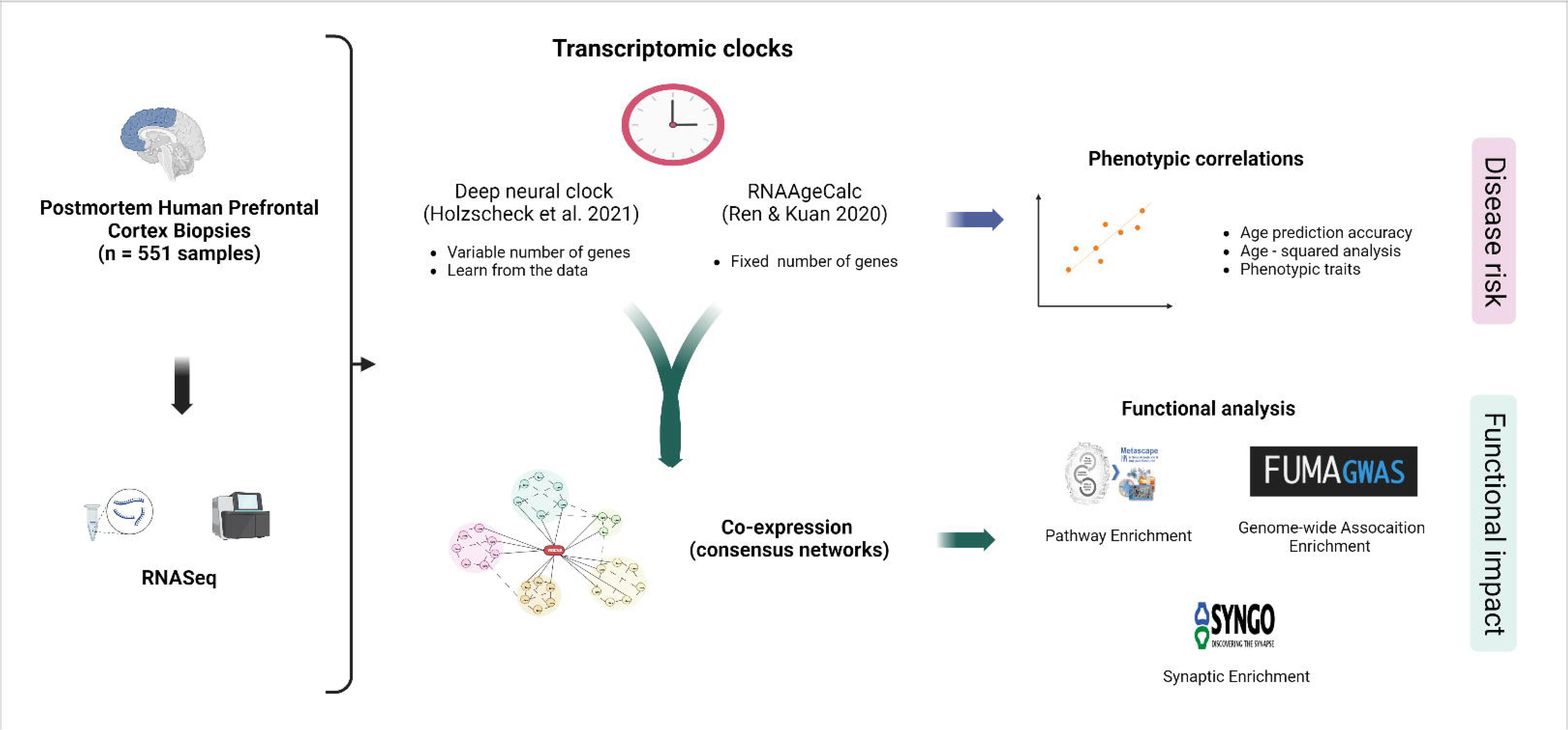
Schematic study workflow. We performed RNAseq of 551 postmortem human prefrontal cortex (PFC) brain samples from 143 individuals and estimated the transcriptomic age using two aging clocks, RNAAgeCalc, and a knowledge-based deep neural network clock (neural clock). Then, we correlated transcriptomic age with age and psychiatric phenotypes and evaluated the prediction accuracy of both clocks. Further, we analyzed the co-expression network shared between the four PFC regions and performed a functional analysis with several databases and algorithms.

## Results

### Transcriptomic clocks performance in predicting age

To estimate the transcriptomic age, we used two machine learning-based transcriptomic clocks, the RNAAgeCalc ^21^ and a clock that uses knowledge-primed artificial neural networks ^22^. The RNAAgeCalc clock is a model that predicts tissue-specific transcriptomic age based on a fixed set of coefficients for the genes calculated from a pre-trained elastic net model. In this analysis, we used the model trained in brain tissue. The knowledge-primed artificial neural network clock (neural clock) is a model that uses the RNAseq data as input and The Molecular Signatures Database (MSigDB) hallmark gene set collection ^45^ to train a neural network and estimate the transcriptomic age. A neural network is divided into layers; each layer has several nodes, and each of these nodes inside a layer is called a neuron. The neuron is relevant to predict the outcome, commonly named activation, if the statistical model associates the neuron with the outcome. Based on the activation of each neuron in the neural network, the model selects a different set of genes as predictors of chronological age. Both transcriptomic clocks showed high correlations with chronological age. The neural clock (r = 0.89, p = 1.89e-56) showed a higher correlation with chronological age compared to the RNAAgeCalc (r = 0.68, p = 3.34e-23) (**Fig. 2A**) (**Supplementary Table 1A**). The clocks also correlated between each other (r = 0.65, p = 2.81e-20) (**Fig. 2B**). The error in age estimation was higher in the RNAAgeCalc (RMSE = 9.09 years) than in the neural clock (RMSE = 5.55 years). Our results suggest that the deep learning clock showed better accuracy in predicting age than RNAAgeCalc.

**Fig. 2.**
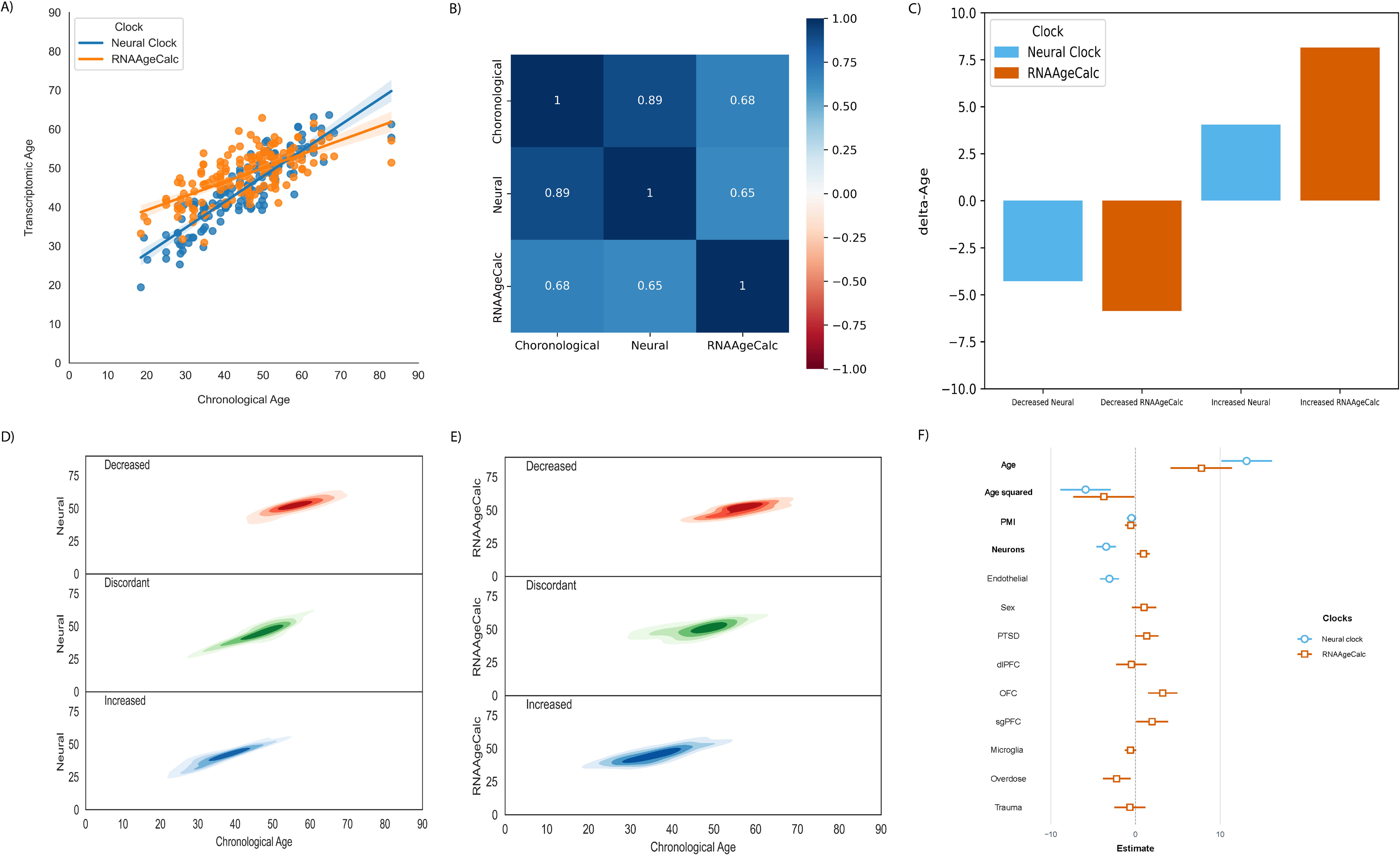
Correlations of transcriptomic age with several phenotypes. A) Correlation between chronological age and transcriptomic age, the orange line represents the RNAAgeCalc and blue the neural clock; B) Pairwise correlations between chronological age and the transcriptomic clocks; C) Mean of the delta of age (dAge) in samples separated by decreased and increased dAge. Blue represents the neural clock, and orange the RNAAgeCalc; D) Density plots of the correlation between chronological and transcriptomic age. Blue represents the density of the distribution of individuals with concordant increased transcriptomic age, green represents samples with discordance between transcriptomic age, red represents concordance with decreased age acceleration, and E) Effect sizes of the most predictive features in the pairwise correlation.

The estimations of delta-Age were higher in the RNAAgeCalc (**Fig. 2C**, **Supplementary Table 1B**). Of the total PFC samples included in the testing set (n = 160), 65% (n = 104) have a concordant delta-Age among both transcriptomic clocks (**Supplementary Table 1C**). We also analyzed the distribution density of the transcriptomic age based on their discordant or concordant effect (increased or decreased) on both clocks (**Fig. 2D** **- 2E**). We found that the density of samples with a concordant decreased delta-Age clustered in advanced ages for both clocks. We also found that samples showing concordant increased transcriptomic age clustered at lower ages, whereas samples showing discordant effects did not show a clustering pattern. These results suggest that transcriptomic clocks have the same prediction accuracy at advanced chronological age, but this prediction accuracy could differ at lower ages. Further, to evaluate if the transcriptomic age could also be correlated with advanced age, we examined the correlation of the transcriptomic age with age and age squared. In the correlation analysis with age squared, we found that the age squared was significant for both clocks (**Supplementary Table 2**). This correlation suggests that the association of transcriptomic age is not linear with chronological age at older age.

### Associations of transcriptomic age with psychiatric-related phenotypes and potential confounding covariates

We analyzed the relationship between transcriptomic age derived from two approaches and psychiatric-related traits, including major depressive disorder, post-traumatic stress disorder, opioid use, alcohol use, tobacco use, amphetamine use, cocaine use, and cause of death and possible confounding variables (PMI, RIN, sex, PFC region, and relative cell-type proportions). First, a multivariate regression analysis was performed followed by a stepwise multivariate regression analysis to refine the variables associated with transcriptomic age. In the multivariate model, we identified multiple associations with each transcriptomic clock (**Supplementary Tables 3A - 4B**). The transcriptomic age estimated with the neural clock was associated with neurons and endothelial cells, relative cell proportion. The RNAAgeCalc transcriptomic age was associated with the OFC and sgPFC. In the stepwise multivariate regression analysis, we identified associations between the Neural clock transcriptomic age and opioid misuse and endothelial cells. In contrast, the RNAAgeCalc was associated with posttraumatic stress disorder (PTSD), orbitofrontal cortex (OFC), and subgenual prefrontal cortex (sgPFC). Concordant associations in both clocks include a negative association with cause of death by overdose and relative neuron proportions (positively associated with RNAgeCalc and negatively associated with the neural clock). Overall, these results suggest that the clocks have differential associations with psychiatric traits.

We also modelled an age squared term in the regression models to account for the effect of increased chronological age in the transcriptomic age. When considering age squared in the model, the age squared term was significant in both clocks (**Supplementary Table 5A - 6B**). In the stepwise correlation analysis, age, age-squared, and neurons remained significant in the Neural clock (**Fig. 2E**), whereas age, age squared, OFC, sgPFC, neurons, and COD overdose remained significant in the RNAAgeCalc. Some of the associations found previously where not replicated when adding the age squared term into the regression models, suggesting that the effect of these traits in the transcriptomic age could not be the same at different chronological ages and considering this variable in the analysis of the PFC transcriptomic age could be important to model aging of the human PFC.

### In silico functional analysis of the genes in the transcriptomic clocks

To investigate potential functional implications of the transcriptomic clocks, we conducted the following analyses: 1) functional enrichment analysis, 2) comparison with a cortical epigenetic clock, and 3) associations with aging phenotypes. The genes used in the RNAAgeCalc and neural clock models differed. These differences rely on the mathematical modeling of the clocks. The RNAAgeCalc uses a fixed number of genes (n = 1616, **Supplementary Table 7**) selected for the association with age, prioritizing each gene by an elastic net algorithm ^21^. The knowledge-primed artificial neural network clock is based on a deep neural network. In this clock, deep neural networks are built by layers where only some layers are activated by the statistical model, to predict an outcome, chronological age. In this clock, each layer is represented by each of the biological pathways in the MSigDB database, building a compartmentalized network. The deep neural clock splits the genes of the transcriptome into each biological pathway, and a gene predicted chronological age if all the genes in this biological pathway are associated with chronological age. This clock selects the best predictors for chronological age in the dataset; in this analysis, the neural clock selected 4325 genes ^22^. Both clocks showed an overlap of 390 genes (7%).

Considering that the genes associated with both clocks could have a greater functional impact on aging-related processes, we performed functional enrichment analysis. By using Metascape, overlapped genes showed enrichment for 976 ontology and pathways (**Supplementary Table 8**). The top enriched ontology/pathways, included diseases of signal transduction by growth factor receptors and second messengers (R-HSA-5663202, ngenes = 45), Epstein-Barr virus infection (hsa05169, ngenes = 27), ribonucleotide metabolic process (GO:0009259, ngenes = 33), Pathways in cancer (hsa05200, ngenes = 36), and cellular response to cytokine stimulus (GO:0071345, ngenes = 40) (**Fig. 3A**). When evaluating previous genome-wide association signals, we identified enrichment in 45 disorders, with diastolic blood pressure, Crohn’s disease, mean corpuscular hemoglobin, and coronary artery disease (**Supplementary Table 9**, **Fig. 3B**) as top associations. Enrichment for mental disorders was also observed, including schizophrenia (ngenes = 26, q-value = 6.84e-04), bipolar disorder (ngenes = 20, q-value = 3.47e-03), and autism spectrum disorder or schizophrenia (ngenes = 20, q-value = 9.88e-03). Overall, these results suggest that the overlapped genes could impact several biological pathways and are associated with aging phenotypes, as well as other health-related traits, including psychiatric traits.

**Fig. 3.**
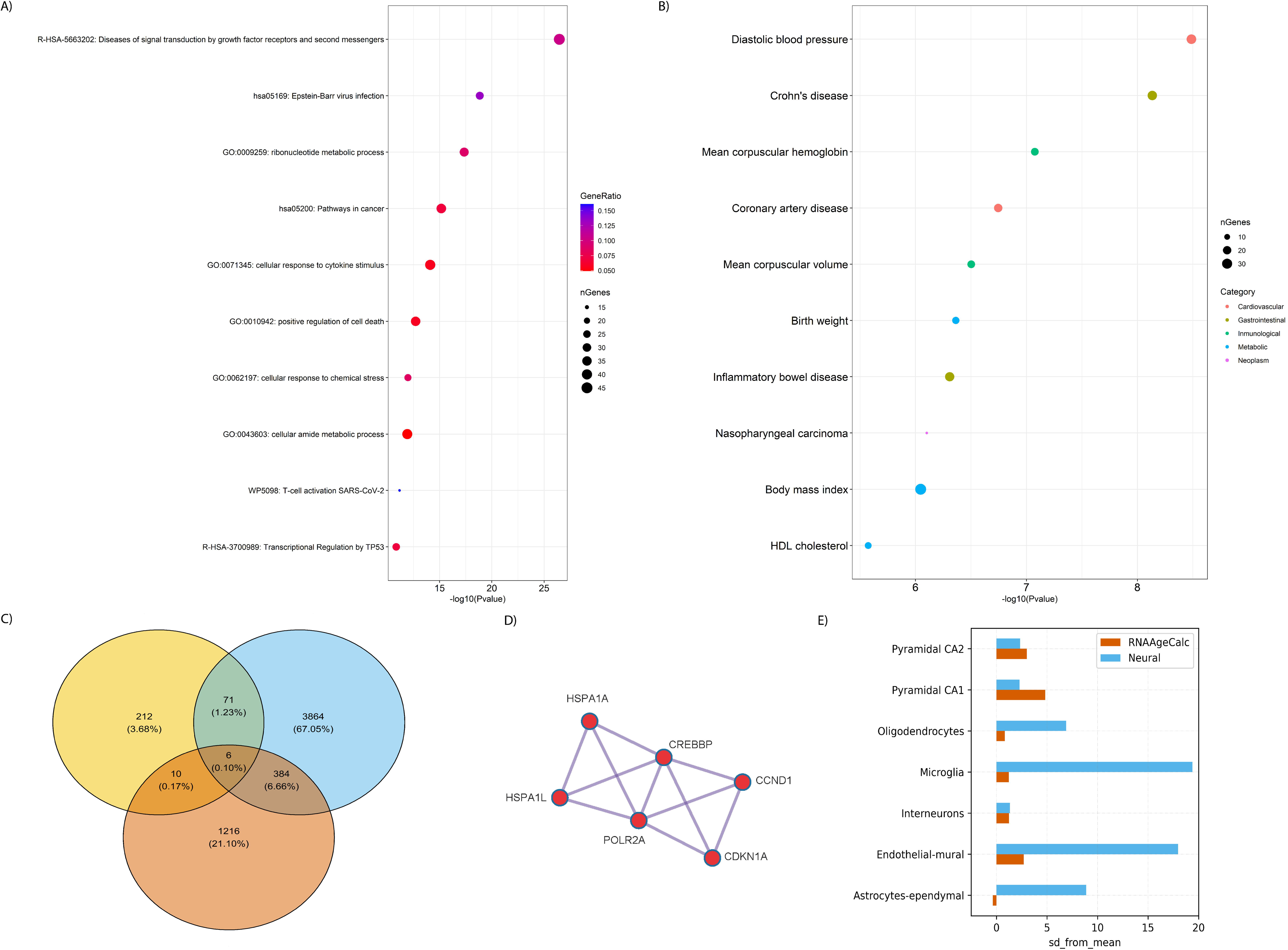
Functional analysis and cell-type enrichment analysis of the overlap of both clocks. A) Top 10 most significant gene ontologies and pathways after redundancy remotion in the clusters; B) Genome-wide enrichment analysis of the genes that overlap between both clocks; C) Gene overlap between the genes used by the transcriptomic locks and the epigenetic clock; D) MCODE of the protein-protein interaction network of the genes that overlap between the neural clock and the epigenetic cortical clock; E) Cell-type enrichment analysis of the genes in the neural clock (blue) and RNAAgeCalc (orange) at annotation level 1.

Epigenetic clocks are the most commonly studied biological clocks and they are good predictors of mortality and health outcomes. We analyzed whether genes of a cortical epigenetic clock trained in the human PFC tissue ^38^ overlapped with the genes of the transcriptomic clocks (**Fig. 3C**). We found six genes overlapping among the three clocks (*POU2F1*, *HLA-C*, *FGF17*, *PVT1*, *ADM*, and *ENO2*). When comparing each transcriptomic clock with the cortical epigenetic clock (**Supplementary Table 10**), we found that the neural clock showed a higher gene overlap (1.20%, ngenes = 72) with the epigenetic clock compared to the RNAAgeCalc (0.20%, ngenes = 10, *RNPC3, GOSR1, ZNF518B, HSF4, TATDN1, TATDN3, HLA-H, POLR1A, NUP62*, and *SSBP4*). The 72 overlapped genes between the neural clock and the epigenetic clock were enriched in 20 ontologies and pathways (**Supplementary Table 10**), with the most significant enrichment including the Nuclear receptors meta-pathway (ngenes = 11) and enzyme-linked receptor protein signaling pathway (ngenes = 12) (**Supplementary Table 11**). In the protein-protein clustering of these 72 genes, performed by Metascape, we identified one gene cluster containing the *HSPA1A*, *HSPA1L*, *CREBBP*, *POLR2A*, *CCND1*, and *CDKN1A* genes (**Fig. 3D**). We evaluated if the genes in both clocks could be regulated by a common transcription factor by enrichment analysis. In the transcription factor enrichment analysis, we identified 55 factors that could target these overlapped genes; the most significant was the *BANP* (**Supplementary Table 12**). We then evaluated the HAGR database ^46^ to identify if any of these transcription factors have been associated with aging phenotypes and, consequently, acting as a regulatory gene in aging processes. We identified *FOXO4*, *E2F1*, *CEBPB*, and *NRF2* with previous associations with aging phenotypes.

### Cell-type enrichment analysis

To understand the cellular diversity associated with each transcriptomic clock, we conducted the Expression Weighted Cell Type Enrichment (EWCE). In the Neural trained clock, we identified enrichment for almost all cells at annotation level 1 (pyramidal CA1, pyramidal SS, astrocytes ependymal, oligodendrocytes, endothelial mural, and microglia) (**Fig. 3E, Supplementary Table 13A - B**). In contrast, RNAAgeCalc was enriched for the pyramidal CA1, pyramidal SS, and endothelial mural cells (**Fig. 3E, Supplementary Table 13C - D**). Thus, these cell-type enrichment analyses suggest that specific cell types differentially contribute to different transcriptomic clocks.

### Consensus network analysis

Gene expression patterns can correlate among genes, and grouping highly correlated genes in networks may identify genes with common biological functions. We used a system biology approach to build a network of the overlapped genes between both clocks followed by functional annotation. We identified 416 nodes and 4855 edges in the consensus network. Of the 390 overlapped genes between both clocks, we identified that 192 genes (49.23%) are co-expressed in all the brain PFC regions (**Supplementary Table 14**). These genes were enriched in 358 ontologies and pathways (**Supplementary Table 15**). The five most enriched pathways were Huntington disease (hsa05016, ngenes = 19), transport of small molecules (R-HSA-382551, ngenes = 26), the citric acid (TCA) cycle/respiratory electron transport (R-HSA-1428517, ngenes = 14), and membrane trafficking (R-HSA-199991, ngenes = 23) (**Fig. 4A**). Also, of the 192 genes identified in the consensus network, we found 71 in the HAGR database (**Supplementary Table 16**). This consensus network showed a high enrichment of protein-protein interactions (PPI) (p-value < 1.00e-06). The PPI network identified 55 genes clustered in 5 groups (**Fig. 4B**). Of the 55 genes identified in these clusters, 5 (*XRCC6, YWHAZ, PRKAR2B, DLD*, and *YWHAG*) were identified in the HAGR database. The genes of this consensus network were also enriched in 14 transcription factors, with *NRF2* as the most significant, a transcription factor also included in the HAGR database (**Fig. 4C**). This consensus network that targets *NRF2* contains 16 genes (*ALDOA*, *ATP1B1*, *ATP6V0A1*, *DMD*, *HSP90AB1*, *PAFAH1B1*, *PFN2*, *PSMD12*, *RAN*, *SKP1*, *TUBA4A*, *YWHAG*, *USP14*, *RTN3*, *CAB39*, and *TUBB*). The consensus network analysis identified genes associated with the aging process and identified *NRF2* as the potential transcription factor regulating the expression of genes common in both clocks.

**Fig. 4.**
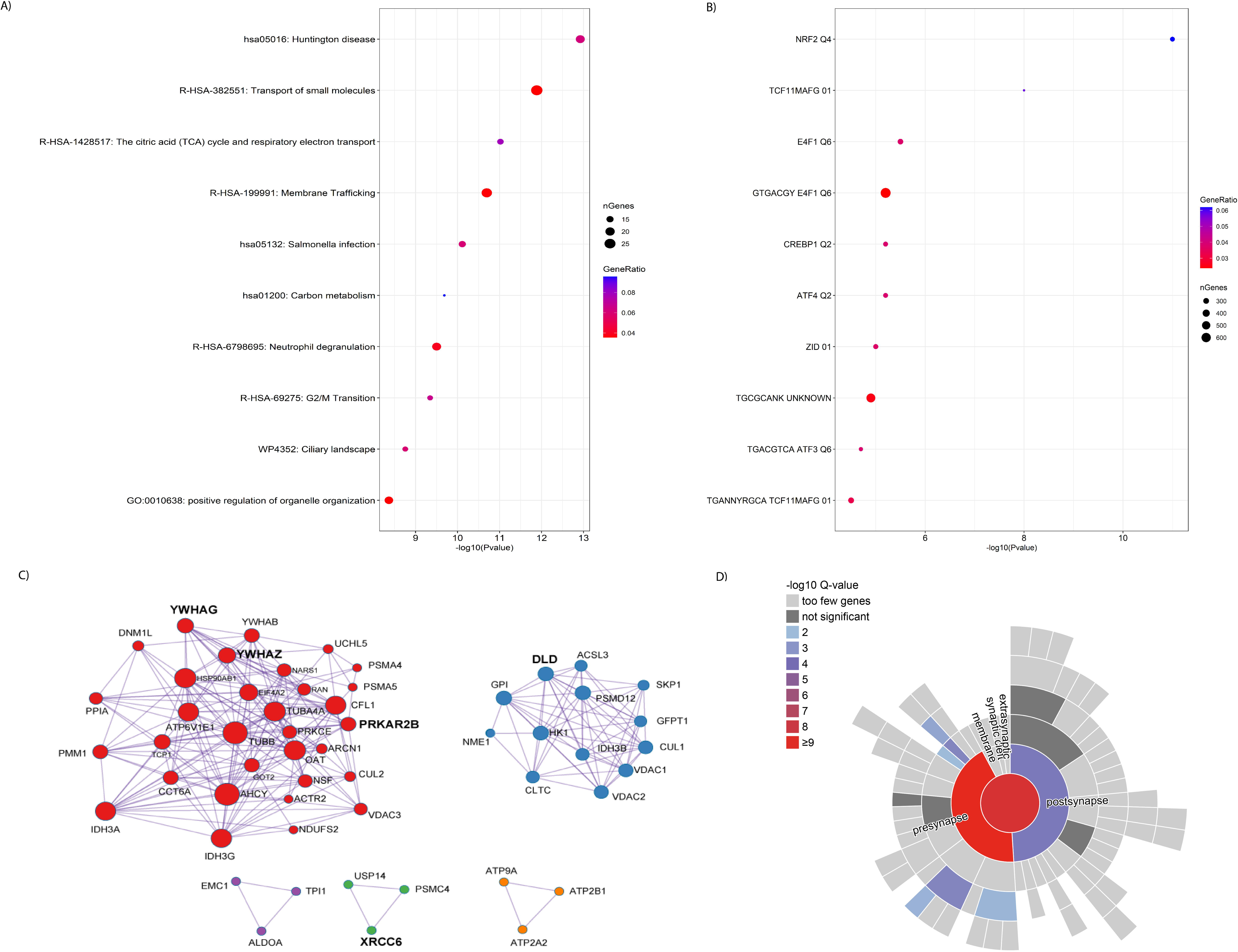
Functional annotation of the consensus network. A) Top 10 enriched gene ontologies and pathways after redundancy remotion; B) MCODE clusters of the protein-protein interaction network analysis; C) Transcription factor enrichment analysis of the consensus network; and D) Synaptic enrichment analysis of the consensus network.

We explored the enrichment of these highly conserved co-expressed genes in the synapsis using SynGO. Enrichment of the consensus network was observed in the synapsis (**Fig. 4D**, **Supplementary Table 17)**, including the *EIF2S1*, *ARPC5L*, *RTN3*, *YWHAG*, *AP2M1*, *NAPA*, *NAPB*, *PFN2*, *HSPA8*, *VPS35*, *ADD2*, *ARFGEF2*, *GAD1*, *PRKCE*, *NSF*, *GDI1*, *VDAC1*, *ATP2B1*, *CLTC*, *DNM1L*, *ATP6V1E1*, *SCAMP1*, *MAL2*, *ATP6V0D1*, *ATP6V0A1*, *CNTN1*, *KCNA4*, *ATP1A3*, *SCN2A*, *PRKAR2B*, *NCKAP1*, *ACTR2*, *YWHAZ*, *DMD*, *PAK3*, and *MAG* genes. Of these 36 genes, six genes (*HSPA8*, *YWHAZ*, *PRKAR2B*, *DMD*, *GAD1*, and *YWHAG*) were found in the HAGR database. This enrichment suggests that genes common in both transcriptomic clocks have a role in synaptic processes.

## Discussion

Here, we report a comprehensive study examining the role and functionality of transcriptomic clocks in a large cohort of human postmortem PFC. Biological clocks, predominantly epigenetic clocks, predict aspects of morbidity and mortality. We compared two different transcriptomic clocks and found that while both can predict age, the knowledge-primed artificial neural networks, trained with deep learning techniques, outperforms RNAAgeCalc. The advantage of deep learning techniques in predicting age has been previously reported for epigenetic clocks ^47,48^. However, even with the higher prediction accuracy, the neural clock correlated with fewer phenotypes compared to the RNAAgeCalc. The observed differences could be due to low gene overlap between the transcriptomic clocks driven by distinct feature selection in each model. This is also observed in epigenetic clocks, where multiple and different sets of CpG sites have been used to predict age, with some achieving comparable accuracy ^49^.

Our stepwise multivariate analysis of RNAAgeCalc found that PTSD is associated with PFC transcriptomic age. This association is consistent with previous findings from peripheral blood and brain. For instance, a previous study using RNAAgeCalc identified an accelerated transcriptional age in 324 World Trade Center responders ^50^. Further, an epigenetic age acceleration in the motor cortex ^51^ and transcriptomic age acceleration in the ventromedial PFC has been reported in PTSD cases^44^. These findings suggest that individuals diagnosed with PTSD exhibit accelerated aging, reflected at both the transcriptomic and epigenomic levels. We also found that the correlation with PTSD was not significant when including an age-squared term in the model, suggesting that the effect of age in transcriptomic age is not linear in the PFC, a phenomenon also observed in epigenetic clocks ^38,52^. A similar effect of the age-squared was found in the correlation of the neural transcriptomic clock and opioid misuse. We found an association of opioid misuse with advanced transcriptomic age, which was not replicated when including the age-squared term in the regression model. This deviation in age at advanced ages could be an effect of the training model. Another explanation is that aging processes could be different at advanced ages compared to other stages of the lifespan. For example, a previous study reported that the effect of body mass index on epigenetic age was found only in young people but not in nonagenarians ^53^. Estimating biological age in older ages is still underexplored; further studies are needed to understand the differences of biological aging across the lifespan.

Our convergent analysis of transcriptomic and epigenetic cortical clocks identified six overlapped genes between the three clocks. These genes are involved in blood-brain barrier integrity and oligodendrocyte function. Among these genes, we found *ADM* (adrenomedullin), a hypotensive peptide secreted by endothelial cells ^54^ and involved in blood-brain barrier structure ^55^. ADM protein is also included in the recently developed epigenetic clock GrimAge ^9^, further supporting its role in aging. Even though the exact mechanisms of *ADM* in aging are still unknown, recent animal studies suggest a role in neurodegenerative diseases ^56,57^. Another convergent gene between the three clocks is *FGF17* (fibroblast growth factor 17), implicated in oligodendrocyte function. Recently, a study where aged mice were infused with cerebrospinal fluid from young mice found that *Fgf17* regulates oligodendrogenesis in the hippocampus, which correlated with an improvement in memory tasks ^58^.

Our consensus network analysis identified 192 co-expressed genes in the four PFC regions shared by both clocks and enriched in synaptic processes, supporting a role of synaptic changes in the PFC during aging ^59^. Further, we identified genes involved in mitochondria functioning and dystrophic neurites. During the aging process, the mitochondria accumulate a plethora of anomalies, promoting an accumulation of dysfunctional mitochondria, which may lead to neuronal alterations and are suggested as common mechanisms for several neurodegenerative diseases, such as Alzheimer’s disease ^60–63^. For example, in patients and animal models of Alzheimer’s disease, imbalance of mitochondrial dynamics ^64^, decreased bioenergetics ^65^, and increased mitochondrial calcium levels are associated with neuronal death ^66^. Besides, the functional analysis of the consensus network found that the transcription factor *NRF2* (Nuclear factor erythroid 2-related factor 2) could be involved in regulating the genes used to predict transcriptomic age. *NRF2* was enriched in the co-expressed genes used as predictors in both clocks. This gene is considered a master regulator of the cellular antioxidant defense system ^67^, controlling the expression of genes with antioxidant response elements (ARE) in their promoters and activating genes with cytoprotective activity ^68–71^. In addition, *NRF2* is upregulated in animal models with exceptional longevity, like the mole-rat ^72,73^, and under caloric restriction, an intervention reported to have an effect in increasing longevity in animal models ^74–78^.

Overall, we conducted a comprehensive transcriptomic clock study of four PFC regions in a large human postmortem brain cohort. Our findings suggest an association between PTSD and transcriptomic aging. Further, we identified functional genes within the human PFC that may play an important role in the aging process.

## Limitations

In the analysis of the knowledge primed deep neural clock, we have to split the sample into a training and testing set, thus reducing our sample size to perform associations with the phenotypic traits. Still, our cohort is larger than similar to recent work ^44^. Also, this reduction in sample size did not allow us to stratify the sample by sex to potential sex-specific effects.

## Methods

### Sample population

We included 551 human postmortem brain samples 143 individuals from the National Center for PTSD Brain Bank ^80^. Psychiatric history and demographic information were obtained by psychological autopsies performed postmortem by next of kin. Diagnostic and Statistical Manual of Mental Disorders version 4 (DSM-IV) criteria and the Structured Clinical Interview for DSM-IV Axis I Disorders (SCID-1) interviews were adapted for psychological autopsy and were used to define the diagnosis. All consents were acquired for the next of kin.

### RNA Sequencing

RNA from each brain region was extracted using the RNeasy Mini kit with genomic DNA elimination, as described by the manufacturer (Qiagen). The RNA integrity number (RIN) and concentration were assessed using a Bioanalyzer (Agilent). Libraries were constructed using a SMARTer Stranded RNA-seq kit (Takara Bio). rRNA depletion was performed using a Ribozero Gold kit (Illumina). Samples were paired-end sequenced on an Illumina HiSeq4000 with a read length of 75 base pairs and targeting a depth of 50 million reads. Sequences in the FASTQ files were mapped to the human genome using STAR (v.2.5.3a) ^81^ with the reference genome (release 79, GRCh38). We used RNA-SeQC ^82^ to quantify the transcripts using Ensembl’s gene transfer format (GTF) annotation file.

### Transcriptomic clocks analyses

To estimate the transcriptomic age, we used two machine learning-based transcriptomic clocks, the RNAAgeCalc ^21^ and a clock that uses knowledge-primed artificial neural networks^22^. The RNAAgeCalc clock is a model that predicts the tissue-specific transcriptomic age based on a fixed set of coefficients for the genes calculated from a pre-trained elastic net model; in this analysis, we used the model trained in brain tissue. We used the *RNAAgeCalc* R package R^21^ and used raw counts of the RNA sequencing analysis from all samples. The knowledge-primmed artificial neural network clock (neural clock) is a model that uses the RNAseq data as input and The Molecular Signatures Database (MSigDB) hallmark gene set collection ^45^ as input to train a neural network and estimate the transcriptomic age. Based on the activation of each neuron in the neural network, the model selects a different set of genes as predictors of chronological age. Conditional quantile normalization was conducted using the *cqn* R package ^83^. For the neural clock analysis, we randomly split the postmortem brain samples into training (⅔ of the brain samples) and testing datasets (⅓ of the brain samples) (**Supplementary Table 18**).

### Predictive age accuracy of transcriptomic clocks

Since the neural clock uses the training dataset to estimate the set of genes that better predicts the chronological age, we only used the testing dataset to explore the predictive accuracy of the clocks. First, we performed pairwise Pearson correlations between the chronological and transcriptomic age; and between the predicted biological ages from both clocks. In addition, we estimated the prediction error of transcriptomic age using the root mean square error (RMSE) measurement implemented in the *rmse* function of the *Metrics* R package. We also calculated the delta of age (delta-Age) as the difference between transcriptomic and chronological age (delta-Age = transcriptomic age - chronological age). If the transcriptomic age is higher than the chronological age (delta-Age > 0), we classified the sample as having an increased transcriptomic age (i.e, positive delta-Age). If the transcriptomic age is lower than the chronological age (delta-Age < 0), we classified the sample as decreased transcriptomic age (i.e, negative delta-Age). To estimate the concordance (i.e, samples with increased transcriptomic age in both clocks) between the delta-Age values of the transcriptomic clocks, we counted the number of samples that had estimated concordance between both clocks and those that had discordant results in both clocks (i.e, samples with increased transcriptomic age in one clock but decreased in the other) and compared these counts by a chi-squared test. All analyses were performed in R version 4.1.1 ^84^.

### Analysis of transcriptomic clocks with age

To test associations between chronological age and transcriptomic age, we regressed transcriptomic age against chronological age. By definition, transcriptomic age is correlated with chronological age, but if it varies non-linearly with chronological age, the estimations in the older ages could be different. Therefore, we tested the extent to which the prediction accuracy of the transcriptomic clocks correlates with age by including an age-squared term in the regression model. All the correlations analyses were performed in R version 4.1.1 ^84^.

### Association of transcriptomic age acceleration with phenotypic traits

To test the association of the transcriptomic clocks with multiple phenotypic traits, we tested multivariable models in two ways: a complete model (a model including all the covariates in the regression model) and a step-wise correlation model to identify the most predictive variables. In this multivariable model, we regressed the transcriptomic age with age, sex, major depressive disorder (MDD), posttraumatic stress disorder (PTSD), opioid misuse, cause of death (classified as overdose, traumatic injury, or other causes of death), tobacco use at the time of death, alcohol use at the time of death, amphetamine misuse, and cocaine misuse. We combined all the brain regions for these models and added the brain region as a covariate. We also included the model’s postmortem interval (PMI), RNA integrity number (RIN), and relative cell-type proportions as possible confounders. Relative cell-type proportions were estimated using the Brain cell type-specific analysis implemented in the *Brettigea* R package ^85^. For the step-wise correlation models, we used the *stepAIC* function in the *MASS* package. We performed the regressions again using the same models, multivariate and step-wise, but including an age-squared variable. The age-squared term was included to verify if the association remained significant after adding the non-linear age variable. All the correlations were analyzed in R ^84^.

### Cell-type enrichment analysis

To understand the cell-type diversity in each transcriptomic clock, we conducted the Expression Weighted Cell Type Enrichment (EWCE) analysis and used the reported single-cell transcriptome reference data. EWCE is a method that uses single-cell transcriptomes to generate the probability distribution of a gene list having an average expression level within different cell types ^86^. This analysis allows us to estimate the enrichment in two levels of cell annotation depending on cell differentiation. For example, annotation level 1 would be interneurons, and annotation level 2 would break this interneuron into 16 different interneurons subtypes depending on your reference dataset.

### Gene overlap with prefrontal cortex epigenetic clock

To identify genes that overlap between transcriptomic and epigenetic clocks, we annotated the 347 CpG sites of a previously published epigenetic clock ^38^, trained to predict epigenetic age in the human prefrontal cortex, to their closest genes using the *IlluminaHumanMethylationEPICanno.ilm10b2.hg19* package.

### Construction of a consensus network (CoDiNA)

Gene co-expression networks are useful for identifying a set of genes that could be biologically relevant and could be coregulated ^87,88^. Most gene co-expression network methods infer independent networks, which could generate inconsistencies in the network construction. The construction of consensus networks has been recently proposed to overcome this limitation ^89^. We used co-expression differential network analysis implemented in the *CoDiNA* package ^90^ to identify a consensus network in the PFC regions. We built independent networks for dACC, OFC, dlPFC, and sgPFC using the *wTO* package ^91^ using the common genes as predictors of age in both transcriptomic clocks. We used conditional quantile normalized counts and extracted the co-expressed genes in the four PFC regions.

### Functional analyses

Functional analyses were conducted, including enrichment analysis and protein-protein interaction (PPI) networks, by using Metascape ^92^, a web-based portal that integrates a broad set of current biological databases and applies analytical pipelines to perform a comprehensive gene list annotation. Most of the known biological databases are redundant, and reducing this redundancy is helpful to have more biological interpretable results. Metascape applies a hierarchical clustering algorithm that reduces redundancies into representative clusters. PPI analysis was performed to infer biologically interpretable results from complex networks. For this, we used Metascape, which applies a mature complex identification algorithm called Molecular Complex Detection (MCODE) ^93^ that detects densely connected regions in PPI networks. Enrichment analyses were performed of the overlapped genes in both transcriptomic clocks and those found in the consensus network.

### Synapsis GO (SynGO) enrichment analysis

Synaptic processes play a role in the brain aging process ^59^. To explore if the transcriptomic clocks include genes with a synaptic function, we performed an enrichment analysis using the GO synapsis database (SynGO) ^94^. We also performed a synapsis GO enrichment analysis of the genes identified in the consensus network.

### Human aging genomic resources (HAGR)

To assess whether the genes in the transcriptomic clocks have been previously associated with aging, we queried the Human aging genomic resource (HAGR) database ^46^, an online collection of multiple databases containing genes associated with aging.

## Data Availability

All data produced in the present study are available upon reasonable request to the authors

## Acknowledgments

We want to thank Dr. Lars Kaderali and Dr. Nicholas Holzscheck for sharing the scripts to model the knowledge-primmed artificial neural network clock. We also want to thank the brain donors and their families thar made this research possible as well as the members of the Traumatic Stress Brain Research Group: Victor E. Alvarez, MD; David Benedek, MD; Alicia Che, PhD; Dianne A. Cruz, MS; David A. Davis, PhD; Ellen Hoffman, MD, PhD; Paul E. Holtzheimer, MD; Bertrand R. Huber, MD, PhD; Alfred Kaye, MD, PhD; Adam T. Labadorf, PhD; Terence M. Keane, PhD; Mark W. Logue, PhD; Ann McKee, MD; Brian Marx, PhD; Deborah Mash, MD; Mark W. Miller, PhD; Crystal Noller, PhD; William K. Scott, PhD; Paula Schnurr, PhD; Thor Stein, MD, PhD; Robert Ursano, MD; Douglas E. Williamson, PhD; Erika J. Wolf, PhD, Keith A. Young, PhD. This work is supported by the U.S. Department of Veterans Affairs via 1IK2CX002095-01A1 (JLMO), the National Center for PTSD, and NIDA R21DA050160 (JLMO).

## Contributions

JLMO and JJMM conceptualized the study. JJMM analyzed the data. JJMM and JLMO wrote the original manuscript. JHK, DLNR, and STN contributed to the interpretation of the results. JHK, MJG, DLNR, STN, and DEAB edited it. All authors revised the manuscript and approved the final version.

## Conflict of interest

No authors report a conflict of interest.

## Notes

### Competing Interest Statement

The authors have declared no competing interest.

### Author Declarations

VA Institutional Review Board.

